# Profiling of circulating glial cells allows accurate blood-based diagnosis of glial malignancies

**DOI:** 10.1101/2022.07.06.22277300

**Authors:** Kevin O’Neill, Nelofer Syed, Timothy Crook, Sudhir Dubey, Mahadev Potharaju, Sewanti Limaye, Anantbhushan Ranade, Giulio Anichini, Vineet Datta

## Abstract

We describe an in vitro test for detection of glial malignancies (GLI-M) based on enrichment and immunostaining of Circulating Glial Cells (CGCs) from peripheral blood sample. Extensive analytical validation studies using U87MG reference cell lines spiked into blood established the analytical performance characteristics of the test. The ability of the test to detect and differentiate GLI-M from non-malignant brain tumors (NBT), non glial type central nervous system (CNS) malignancies (NGCM), brain metastases from primary epithelial malignancies in other organs and healthy individuals were evaluated in four studies. The cumulative performance metrics of the test across all 4 clinical studies were 99.35% Sensitivity (95%CI: 96.44% - 99.98%) and 100% Specificity (95%CI: 99.37% - 100%). The performance characteristics of this test support its clinical utility for diagnostic triaging of individuals presenting with ICSOL.

## INTRODUCTION

Brain tumors account for 85% to 90% of all primary central nervous system (CNS) tumors (1) as well as ∼300,000 (∼1.6%) of the total ∼19,300,000 annual cancer incidences and 250,000 (∼2.5%) of the total 10,000,000 annual cancer-related mortality globally (2). In patients presenting with radiological intracranial space occupying lesions (ICSOL), the differential diagnosis includes primary glial malignancy (GLI-M) and metastases from other solid tumors. Non-malignant ICSOL are more common (3) than GLI-M and have different management, emphasising the critical importance of expeditious establishment of diagnosis. Glioblastome multiforme (GBM) is the most common (49%) subtype of all malignant tumors (3).

Standard of care (SoC) for establishing the diagnosis in such individuals presenting with ICSOL is histopathological evaluation (HPE) of tumor tissue specimens obtained from surgical excision or biopsy. Surgical resection or biopsy are more challenging under circumstances of poor patient performance, in the presence of comorbidities or patients’ reluctance (4). Procedural risks are well-documented and include pain and discomfort, intracranial haemorrhage, cerebral edema, infections as well as morbidity and mortality (5). Furthermore, the anatomical site of the lesion may be associated with increased procedural risks and complications. prior studies also suggest that around 70% of patients with intracranial lesions have benign conditions (3) indicating that in a sizeable population of symptomatic individuals, the ability to obtain the same inference non-invasively would significantly reduce the requirement for invasive biopsy.

There is therefore considerable benefit in non-invasive detection of GLI-M including risk mitigation, resource optimization, cost benefits and avoidance of delays in time to diagnosis and time to treatment, especially in unresectable cases where tissue sampling is unviable. Previous attempts at non-invasive detection of GLI-M and at differentiating GLI-M from non-malignant brain tumors (NBT) and brain metastases have examined profiling of gene variants (6) or CpG island methylation (7) in cell-free DNA and profiling of exosomal mRNA / miRNA transcripts (8). However, these approaches have been limited by lower sensitivity and specificity (9). Circulating tumor cells (CTCs) are viable tumor derived cells in circulation, the molecular evaluation of which may be an alternative to or comparable with that of the tumor tissue from which they originate (10-12). CTCs are rarely detected in the peripheral blood of healthy individuals and their detection in such populations may be an indication of an underlying malignancy (13, 14).

We have previously described functional enrichment of CTCs and CGCs from peripheral blood using a proprietary CGC/CTC enrichment medium (CEM) which selectively induces apoptosis in non-malignant cells and permits survival of malignant cells. This method yields sufficient viable CGCs/CTCs for downstream applications including multiplexed immunocytochemistry (ICC) (13,15). In the present study we have used this enrichment method for harvesting CGCs from blood samples of patients with GLI-M and identification based on co-expression of GFAP and OLIG-2 as determined by ICC profiling of the harvested CGCs. We describe the performance characteristics of the blood-based test to detect GLI-M and differentiate it from NBTand EPI-M with brain metastases.

## RESULTS

### Method Feasibility

#### Detection Thresholds

The fluorescence intensity (FI) of GFAP and OLIG2 were higher in U87MG, pooled circulating glial cells (CGCs) and malignant (glial) tumor derived cells (M-TDCs) than in MOLT-3 or low-grade / non-malignant brain tumor derived cells (B-TDCs). The FI of CD45 was higher in MOLT-3 than in U87MG, CGCs, M-TDCs or B-TDCs (Supplementary Figure S1). Based on these findings, the FI threshold for CGC positivity was assigned as 42,000 (relative fluorescence units, RFU) for GFAP and 56,000 for OLIG2; these also apply as a lower threshold for U87MG, which is the positive control (PC) for both markers. For CD45 36,000 RFU was set as the upper threshold in U87MG, M-TDC and CGCs where CD45 expression is not expected; it is also the lower threshold for MOLT3 which is the PC for CD45 and negative control (NC) for all other markers.

#### Marker Specificity

As seen in Supplementary Figure 2, CTCs from various (non-CNS) cancer types had low expression (FI) of GFAP and OLIG2.

#### Marker Expression in Glial Malignancies

As seen in Supplementary Figures S3 and S4, the expression (FI) of GFAP and OLIG2 were determined to be above the positivity thresholds when evaluated on CGCs from various subsets of patients stratified by gender and age-group (n = 34) or histological subtype and grade (n = 63). Thus none of these factors contributed to loss of detection sensitivity of the test.

### Analytical Validation

Table 2 is a summary of all the findings of the analytical validation study.

**Table 1.**
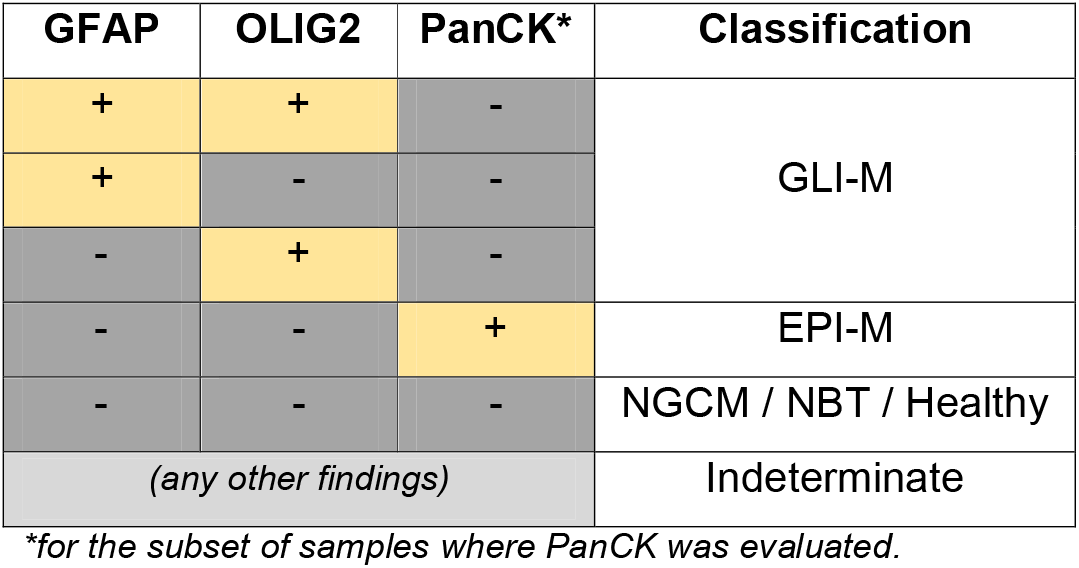
Decision Matrix. The Decision Matrix shows the relation between various marker expression combinations and the predictions for each sample. Primary classification of samples is based on expression of GFAP and OLIG2. The matrix also explains the classification in a subset of samples where PanCK was used in addition to the above markers. GLI-M: Glial malignancy; NGCM: Non-glial central nervous system malignancy; NBT: non-malignant brain tumor; EPI-M: epithelial malignancy with brain metastases.

**Table 2.** Summary of Analytical Validation. The findings of Analytical Validation indicate that the Test provides consistent, accurate and reproducible results with little or no interference from routine endogenous or exogenous factors when samples are obtained, stored and processed under the recommended conditions. Comprehensive details are provided in Supplementary materials. Numbers within parentheses indicate 95% confidence intervals (95% CI). The two values for repeatability and within laboratory precision are the CV (%) at 5 cells / 5 mL (positivity threshold) and 15 cells / 5 mL (3× positivity threshold) respectively.

| **Parameter** | **Value** |
| --- | --- |
| **Analyte Stability** | 48 h |
| **Linearity** | $R^2 > 0.99$ |
| **Linearity Interval** | 5 – 1280 cells / 5 mL |
| **Limit of Blank** | 0 cells / 5 mL |
| **Limit of Detection** | 1 cell / 5 mL |
| **Limit of Quantitation** | 6 cells / 5 mL |
| **Specificity** | 100% (85.8% - 100%) |
| **Sensitivity** | 95.0% (83.1% - 99.4%) |
| **Accuracy** | 96.9% (89.2% - 99.6%) |
| **Repeatability** | 13.7% (at 5 cells / 5 mL),<br>10.0% (at 15 cells / 5 mL) |
| **Within Laboratory Precision** | 23.5% (at 5 cells / 5 mL),<br>13.7% (at 15 cells / 5 mL) |

#### Stability and Recovery

In the spiked samples, ≥80% recovery was observed for each cell type for up to 48 h (Supplementary Table S1). In clinical samples, the CTC recovery was ≥85% at 48 h, when 0 h recovery was normalized as 100%. The findings of the stability and recovery studies indicated that clinical samples could be stored at 2°C-8°C for up to 48 h with ≤15% loss of cells.

#### Linearity

The linearity interval was determined to be 5 - 1280 cells / 5 mL based on lower limit of linear interval (LLLI) being 5 cells / 5 mL and upper limit of linear interval (ULLI) being 1280 cells / 5 mL for both markers. Similarly, R^2^ ≥0.99 for both markers demonstrated the linear response characteristics of the method (Supplementary Figure S5, Supplementary Table S2). At the sample positivigy threshold of 5 cells / 5 mL, the observed deviation from linearity was -17% for GFAP and -19% for OLIG2, which are within the permissible range of -26% to +22% for 15% ADL, as specified in CLSI EP06.

#### Limits of Blank, Detection and Quantitation

LoB, LoD and LoQ were determined as per CLSI recommended guideline EP17-A2. No (marker positive) cells were detected in any of the unspiked samples (no false positives). Thus, the limit of blank was determined to be 0 cells / mL. The Limit of Detection (LoD) was determined as 1 cell / 5 mL for both markers. For the LoQ, the Allowable Deviation from Linearity (ADL) was pre-specified at 15%. The LoQ was determined to be 6 cells / 5 mL for GFAP and 5 cells / 5 mL for OLIG2, thus the overall LoQ was 6 cells / 5 mL.

#### Sensitivity, Specificity and Accuracy

Based on recovery of marker positive cells in 40 spiked samples (5 – 80 cells / 5 mL), the sensitivity was 92.5% for GFAP and 95.0% for OLIG2. Since marker positive cells were undetectable in any of the un-spiked samples (per marker), the specificity was deemed to be 100%. Accuracy was determined to be 95.3% for GFAP and 96.9% for OLIG2 (Supplementary Table S3).

#### Precision

Supplementary Table S4 provides the observed mean, SD and CV(%) along with the 95% CI for repeatability and within laboratory precision.. The %CVs were 13,7% for repeatability and 23,5% for within laboratory precision at the detection threshold and 10,0% for repeatability and 13,7% for within laboratory precision at 3× the detection threshold. The higher observed CV at the lower spike density is a typical and expected feature of in vitro detection tests.

#### Interfering Substances

The presence of drugs at medically relevant peak plasma concentrations (C_Max_) or the deranged (clinically, high) serum parameters did not significantly impact the sensitivity of the test for detection of spiked U87MG cells (Supplementary Table S5). The study established the ability of the test to remain unaffected in presence of systemic treatment agents (drugs) and elevated serum parameters.

### Clinical Study Findings

The ability of the test to detect and differentiate samples from GLI-M and NBT was first established in a stringent, blinded cross-validation study which was designed to minimize the risk of overfitting in the training set. The inclusion criteria are provided in Supplementary Table S6 and the demographics of this cohort are provided in Supplementary Table S7. The observations in the training and test set samples are provided in Supplementary Table S8. Among the 101 GLI-M cases in the Training Set, 100 were positive (99%) and 1 was negative (1%) for CGCs. Among the 31 cases of NBT, 1 (3.2%) was positive for CGCs and 30 (96.8%) were negative for CGCs. In absence of follow-up data demonstrating diagnosis of GLI-M, the positive NBT case was considered as a false positive. In the Test Set (n = 57), there were 44 samples with positive and 13 samples with negative findings. All 44 positive samples were determined to be GLI-M yielding a sensitivity of 100%. All negative samples were determined to be NBT yielding a specificity of 100% (Supplementary Table S9).

The ability of the test to detect and differentiate samples from GLI-M, NBT, epithelial malignancies with brain metastatses (EPI-M) and healthy individuals was next established in a second stringent, blinded study. The inclusion criteria are provided in Supplementary Table S10 and demographics of this cohort are provided in Supplementary Table S11. The observations in the study samples and the performance characteristics are provided in Supplementary Table S12. Among the 40 GLI-M samples, none were positive for CTCs while all were positive for CGCs. Among the 24 EPI-M samples, none were positive for CGCs and all were positive for CTCs. Among the samples fromNBT cases (n = 22) and healthy individuals (n = 500), none were positive for CGCs or CTCs. The test thus has a CGC detection sensitivity of 100% and specificity of 100% for detection of CGCs.

The ability of the test to identify and differentiate GLI-M and NBT was evaluated in a prospective multi-centric study cohort of 68 patients presenting with ICSOL. The inclusion criteria are provided in Supplementary Table S13 and the demographics of the cohort in Supplementary Table S14. The observations on samples (status of CGCs) are summarized in Supplementary Table S15. Of the 68 cases, 56 were positive for CGCs and 12 were negative as per the Decision Matrix. After unblinding, it was revealed that all 56 positive samples were GLI-M and all 12 samples were NBT. The test had a sensitivity of 100% as well as a specificity of 100% for detection of GLI-M and differentiating GLI-M from NBT(Supplementary Table S16).

The ability of the test to identify and differentiate GLI-M from NBT and non-glial central nervous system (CNS) malignancies (NGCM) was evaluated in a prospective cohort of 31 patients presenting with intra-axial ICSOL. The inclusion criteria are provided in Supplementary Table S17 and the demographics in Supplementary Table S18. Of the 31 cases, 13 were positive for CGCs and 18 were negative. After unblinding, it was revealed that all 13 positive samples were GLI-M. Of the 18 negative samples, 1 was GLI-M, 8 were NGCM and 9 were NBT (Supplementary Table S19). The test thus had a sensitivity of 92.9% as well as a specificity of 100% for detection of GLI-M and differentiating GLI-M from NBT (Supplementary Table S20).

Table 3 provides a summary of the study-wise performance characteristics as well as the cumulative performance characteristics.

**Table 3.** Clinical Performance Characteristics of the Test. Performance characteristics of the test were determined in four separate clinical studies.. In all studies, the sensitivity, specificity and accuracy were determined. In the test design, samples with equivocal findings for GLI-M are considered as positive and recommended for clinical follow-up. Thus the performance characteristics were determined with the same consideration.

|  | **Sensitivity** | **Specificity** | **Accuracy** |
| --- | --- | --- | --- |
| **Clinical Study-1**<br>**(Validation Set)** | 100%<br>(95% CI: 91.96% - 100%)<br>(n = 44) | 100%<br>(95% CI: 75.29% - 100%)<br>(n = 13) | 100%<br>(95% CI: 93.73% - 100%)<br>(n = 57) |
| **Clinical Study-2** | 100%<br>(95%CI: 91.19% - 100%)<br>(n = 40) | 100%<br>(95%CI: 99.33% - 100%)<br>(n = 546) | 100%<br>(95%CI: 99.37% - 100%)<br>(n = 586) |
| **Clinical Study-3** | 100%<br>(95%CI: 93.62% - 100%)<br>(n = 56) | 100%<br>(95%CI: 73.54% - 100%)<br>(n = 12) | 100%<br>(95%CI: 94.72% - 100%)<br>(n = 68) |
| **Clinical Study-4** | 92.86%<br>(95%CI: 66.13% - 99.82%)<br>(n = 14) | 100%<br>(95%CI: 80.49% - 100%)<br>(n = 17) | 96.77%<br>(95%CI: 83.30% - 99.92%)<br>(n = 31) |
| **Cumulative** | 99.35%<br>(95%CI: 96.44% - 99.98%)<br>(n = 154) | 100%<br>(95%CI: 99.37% - 100%)<br>(n = 588) | 99.87%<br>(95%CI: 99.25% - 100%)<br>(n = 742) |

### Orthogonal Verification – FISH

Among the 22 cases of NBT, there were no instances of EGFR copy gain detected by FISH on tumor tissue. All 22 samples were also negative for CGCs indicating 100% concordance for specificity (Malignant v/s Benign). Among the 22 cases of GLI-M, EGFR copy gain was observed on tumor tissue in 8 cases, all of which were also detectable on CGCs indicating 100% concordance (sensitivity). Among the remaining 14 samples with normal EGFR status by FISH, the CGCs also showed normal status indicating 100% concordance (specificity).

## DISCUSSION

Presentation of patients with intra-cranial malignancy is frequently symptomatically non-specific and differentiating such patients from those with non-malignant conditions or with absent pathology is challenging. Indicative of this, GBM presents as a medical emergency more frequently than any other common cancer, implying that effective strategies for rapid diagnostic stratification of patients presenting with suspicious symptoms are urgently required. Furthermore, it is clearly critical to differentiate GLI-M from NBT or metastases from other solid tumors. Obtaining a tissue diagnosis via biopsy of ICSOL is often challenging and has well-described risks.

Here we describe a blood-based test for detection of GLI-M in individuals presenting with ICSOL, based on detection of CGCs by multiplexed fluorescence ICC profiling. The test can detect common subtypes that account for about 97% of all GLI-M, irrespective of age, gender, subtype and grade. The analytical validation of our platform confirmed accuracy and reliability of the test. The clinical validation study demonstrated overall >99% sensitivity and 100% specificity for detection of GLI-M. The performance characteristics of the test favour clinical adoption of this technology for supporting more effective diagnosis in individuals presenting with ICSOL, especially among patients with unresectable or non-biopsiable ICSOLs. To our knowledge, there are presently no non-invasive or non-radiological tests for detection of GLI-M in individuals with ICSOLs.

Our test is based on the detection of circulating tumor cells (CTCs), which in the context of a glial malignancy are called circulating gial cells (CGCs). In primary solid organ cancers, the existence of CTCs is linked to dissemination and metastatic spread. Extracranial metastases though rare in GLI-M, have been reported previously (16-20). The detection of circulating (malignant) glial cells (CGCs) in blood samples from patients with GLI-M appears to indicate that while CGCs can enter circulation, they may be unable to find a target tissue where they can egress, survive, and grow (21). Zhang et al hypothesize that the inability to detect extracranial metastasis may be a consequence of the low survival (shorter life span) of patients with GLI-M, and that the probability of detecting extracranial metastases may be higher in patients who survive longer (22).

Prior studies have shown the presence of CGCs in low and high grade gliomas and glioblastomas, as well as their absence in healthy individuals and those with non-malignant brain tumors.Bang-Christensen et al reported 0.5 – 42 CGCs / 3 mL blood irrespective of grade or subtype of GLI-M (23) via a novel immunocapture method. MacArthur et al used density-gradient centrifugation followed by telomerase assay and Nestin expression to detect CGCs in 8 out of 11 (72%) cases of radiation-naïve glioma with an average of 8.8 CGCs / mL of blood (10). Sullivan et al demonstrated that mesenchymal like properties of CGCs could contribute to their invasiveness, allowing them to enter into circulation (24). Based on chromosome 8 polyploidy and immunostaining for GFAP (positive) and CD45 (negative), Gao et al reported CGCs in peripheral blood of 24 out of 31 (77%) patients with GLI-M with no correlation between the number of CGCs and the subtype / grade of malignancy (25). Similarly, Krol et al reported CGC clusters in 7 of 13 (53.8%) cases of glioblastoma (26).

Our test detects CGCs based cellular GFAP and OLIG-2 expression and provides unambiguous evidence of the underlying malignancy in the form of directly visualized malignant cells. The detection of GFAP and OLIG2 positivity on malignant cells is not prone to confounding, as may be observed in case of various serum cancer antigens which are often elevated in patients with non-malignant conditions. Positive marker expression on cells is based on standardized fluorescence intensities (FI) detected using a sensitive and automated high content screening platform which minimizes the risk of false negatives. Our test showed high sensitivity and specificity for CGCs detection in analytical validations as well as in the clinical studies.

Our study shows that it is possible to obtain sufficient viable CGCs in peripheral blood samples for detection of GLI-M and differentiation of GLI-M from NBT and brain metastases of solid tumors. To our knowledge, the test described in this manuscript is the first of its kind which uses a hallmark property of malignancy for enrichment of CGCs. Applications of this core technology for detection of breast and prostate cancers have been previously described (27,28). Our test is minimally invasive and is performed on a peripheral (venous) blood draw. The test is not currently intended to replace standard diagnostic imaging or tissue sampling. Contemporary brain imaging such as magnetic resonance imaging (MRI) offers diagnostic guidance in ICSOLs by being able to distinguish some malignant and non-malignant ICSOLs based on radiological morphology. We envisage the test to provide an additional layer of high quality evidence which can potentially support diagnostic and disease management decisions prior to lifting the scalpel. The CGC-based approach described in this study may be especially relevant in cases with unresectable or non-biopsiable ICSOLs which can pose a diagnostic roadblock. Surgical resection may not be viable due to the proximity of the lesion to regions associated with vital functions or comorbidities; up to 40% of cases with advanced or high-grade brain lesions are reported to be unresectable (29). Further, brain biopsies have been reported to be unviable, inconclusive, or non-diagnostic in up to 20% of cases (30-32)

In such cases, the test findings have the potential to mitigate any risks of overtreatment in individuals with non-malignant brain tumors as well as the potential to reduce risks associated with delayed diagnosis and treatment in individuals with GLI-M. In addition, the detection and differentiation of CGCs and (epithelial) CTCs based on marker expression profile can also aid the differentiation of primary CNS (glial) malignancy and brain metastases of a solid tumor from a non-CNS primary; albeit rarer, there are reports of cancers presenting with brain metastases [33] and such cases represent yet another subset of patients who may benefit from our approach. The strength of our study is the multiple clinical studies with blinded sample analysis, all of which demonstrated high concordance between test findings and clinical diagnosis and support clinical application of the test.

The high clinical sensitivity indicates a very low risk of missing GLI-M while the high specificity indicates an imperceptible (if any) risk of false positive findings in individuals without a primary GLI-M. Although the assay has high performance characteristics for detection of GLI-M, the test does not detect rarer subtypes such as CNS lymphoma and gliosarcoma. The test is also not intended to differentiate the subtype or grade of malignancy. The 2021 World Health Organization (WHO) guidance for classification of CNS tumors (34) emphasizes on the increasing role of molecular diagnosis by considering gene variants as prognostic features. Advancements in next generation sequencing (NGS) technology platforms suggest a potential for molecular profiling of glial malignancies using the limited yields of tumor nucleic acids (TNA) isolated from CGCs (35). Thus, while the test is currently not intended to replace standard tissue sampling, we envisage future iterations of our test to include immune-profiling of CGCs as well as molecular profiling of CGC-derived TNA for a more holistic role in diagnostic work-up with reduced dependence on tumor tissue.

The approach described in this manuscript requires only a peripheral blood draw from the patients. The simplicity of a blood based test makes it amenable to integration within existing standard of care diagnostic pathways in most healthcare systems. Blood collection is a simple, low risk procedure that can be performed at any primary healthcare centre or physician’s clinic or even a pharmacy. From the patient’s perspective, there are no additional visits to advanced healthcare facilities or additional wait times. From the healthcare provider’s point of view, no additional resources or infrastructural investments are required.

In conclusion, we present a blood-based, non-radiological test for detection of glial malignancies with potential clinical applications in symptomatic individuals who are advised an invasive biopsy as part of standard diagnostic work-up as well as diagnostic support in individuals with unresectable and / or non-biopsiable ICSOLs. Our test has potential to enable more effective clinical decision making by providing direct evidence of the presence of GLI-M in these cases.

## ONLINE METHODS

### Patients and Samples

All biological samples reported in this manuscript were primarily obtained from participants in four clinical studies to identify blood-biomarkers for detection of various types of malignancies and to differentiate cancer cases from individuals with benign conditions or healthy individuals.

All biological samples were obtained from participants in four studies,

1. GlioLENS with CTRI number CTRI/2019/02/017663

(http://ctri.nic.in/Clinicaltrials/pmaindet2.php?trialid=31387),

2. TRUEBLOOD with CTRI number CTRI/2019/03/017918

(http://ctri.nic.in/Clinicaltrials/pmaindet2.php?trialid=31879) and

3. RESOLUTE with CTRI number CTRI/2019/01/017219

(http://ctri.nic.in/Clinicaltrials/pmaindet2.php?trialid=30733).

4. Prospective Study at The Imperial College, London

(this study has not been registered at any clinical trial repository)

The Institutional Ethics Committee (IEC) of Datar Cancer Genetics (DCG, sponsor) gave ethical approval for GlioLENS, TRUEBLOOD and RESOLUTE. IEC – Clinical Studies of Apollo Hospitals, Hyderabad gave ethical approval for GlioLENS. The Imperial College Ethics Committee / Institutional Review Board gave ethical approval for the prospective study conducted at Imperial College, London.

The GlioLENS study enrolled known cases of GLI-M as well as symptomatic individuals with ICSOL suspected of GLI-M. The TRUEBLOOD study enrolled known cases of cancer and symptomatic individuals with suspected solid organ cancers and the RESOLUTE study enrolled asymptomatic adults with no prior diagnosis of cancer and no current symptoms or clinical features of cancer. Imperial College study enrolled surgery- and biopsy-naïve adults with ICSOLs to determine concordance between the detection of CGCs in pre-surgery / pre-biopsy blood and subsequent HPE diagnosis on tumor tissue.

All studies were performed in accordance with the Declaration of Helsinki as well as any applicable regulatory guidelines. Written informed consent was obtained from adult study participants or their parents in case of patients aged less than 18 years. All studies were performed in accordance with the Declaration of Helsinki. Specimens were processed at the CAP and CLIA accredited facilities of the sponsor, which also adhere to quality standards ISO 9001:2015, ISO 27001:2013 and ISO 15189:2012. Fifteen millilitres of peripheral blood was collected from all adult study participants in EDTA vacutainers. In 3 patients aged less than 10 years, 5 mL blood was collected and in patients aged 10 – 17 years, 10 mL blood was collected. For suspected cases of GLI-M, blood collection was performed prior to the patients undergoing tissue sampling or neurosurgical excision, and where possible leftover tissue samples were also obtained from consenting participants. Biological samples were stored at 2°C – 8°C in appropriate tumor transport media during transport to reach the clinical laboratory within 48 h. The status of all samples was blinded to the operators (those who performed the enrichment and ICC) as well as the analysts (those who analyzed the data) by assigning unique 10-digit alphanumeric barcodes to minimize potential biases arising from prior knowledge of sample status. The reporting of observational studies in this manuscript is compliant with STROBE guidelines (36).

### Antisera, Reference Cells and Reagents

The antisera used included recombinant human (RH) Anti-CD45-IgG-APCVio 770 (Miltenyi Biotec), Mouse Anti-GFAP IgG (Biocare Medical), Mouse Anti-OLIG2 IgG (Cell Marque), Anti-mouse Alexa Fluor 594 (Invitrogen), Anti-mouse Alexa Fluor 488 (Invitrogen) and RH Anti-CK-IgG1-Vio 515 (Miltenyi Biotech). U87MG (glioblastoma) was used as the reference cell line for spiking studies for analytical validation. MOLT-3 (leukemia). All cell lines were procured from American Type Culture Collection (ATCC) directly or from other licensed (authorized) vendors. The identity of reference cell lines was confirmed periodically by Short Tandem Repeat (STR) Profiling as per existing quality standards at the sponsor’s facilities and by using a commercial kit as per the manufacturer’s recommendations. The cell lines were also periodically tested for Mycoplasma as per existing quality standards at the sponsor’s facilities and by using a commercial kit as per the manufacturer’s recommendations. All reagents described in the manuscript are cell culture or molecular biology grade and have been procured from licensed commercial vendors.

### Enrichment of Circulating Tumor/Glial Cells from Peripheral Blood

Blood samples (5 mL or 7.5 mL) were processed for the enrichment of CGCs/CTCs from peripheral blood mononuclear cells (PBMC) as described previously (13, 15). Briefly, PBMCs were isolated from whole blood via lysis of red blood cells (RBCs) followed by centrifugation. PBMCs resuspended in Phosphate Buffered Saline (PBS) were treated with CEM for 5 days at 37°C, 5% CO_2_, 65% relative humidity. During treatment, the CEM induces cell death in all non-malignant (hemato-lymphoid, epithelial and endothelial) cells, while malignant tumor derived cells (CTCs) survive. On the firth day, surviving cells and cell clusters are harvested by centrifugation and resuspended in PBS.

### Isolation of Primary Tumor Derived Cells

The isolation of primary tumor derived cells (TDCs) from an excised tumor (malignant / benign) has been described previously (37).

### Immunocytochemistry Profiling of Cells

Immunocytochemistry (ICC) profiling of samples for identification of CTCs/CGCs was performed as described previously (15). Briefly, viable apoptosis-resistant cells enriched from 5 mL of blood were resuspended in 1000 μL 1x Phosphate Buffered Saline (PBS) and 100 μL aliquots were seeded into 10 wells (each well is equivalent to 500 μL blood sample). Cells were fixed with 4% paraformaldehyde, permeabilized with 0.3% Triton-X 100 and treated with 3% BSA (blocking). Cells in separate wells were immunostained with separate primary (1°) Ab cocktails for multiplexed analysis of marker combinations to identify CGCs, (a) 1:100 Anti-GFAP + 1:500 Anti-CD45, and (b) 1:100 Anti-OLIG2 + 1:500 Anti-CD45. In the subset of samples where cells were enriched from 7.5 mL blood, the status of a third marker combination, i.e., 1:500 Anti-PanCK + 1:500 Anti-CD45 was evaluated in addition to the above to identify CTCs from epithelial malignancies (EPI-M). Each marker combination was evaluated in 5 wells (500 μL × 5 = 2.5 mL equivalent of blood). Samples were washed with PBS and incubated with respective secondary (2°) Ab. Finally, cells were washed with PBS and treated with 4’,6-Diamidino-2-phenylindole dihydrochloride (DAPI) for nuclear staining. Control samples were included in each run. Samples were evaluated on the Cell-Insight CX7 High Content Screening (HCS) Platform (Thermo Fisher Scientific, Waltham, MA, USA) to determine the Fluorescence Intensity (FI) for each marker. Figure 1 is a schema of the functional enrichment of malignant cells and ICC profiling to detect CGCs. The decision matrix for assigning samples into various categories is provided in Table 1.

**Figure 1.**
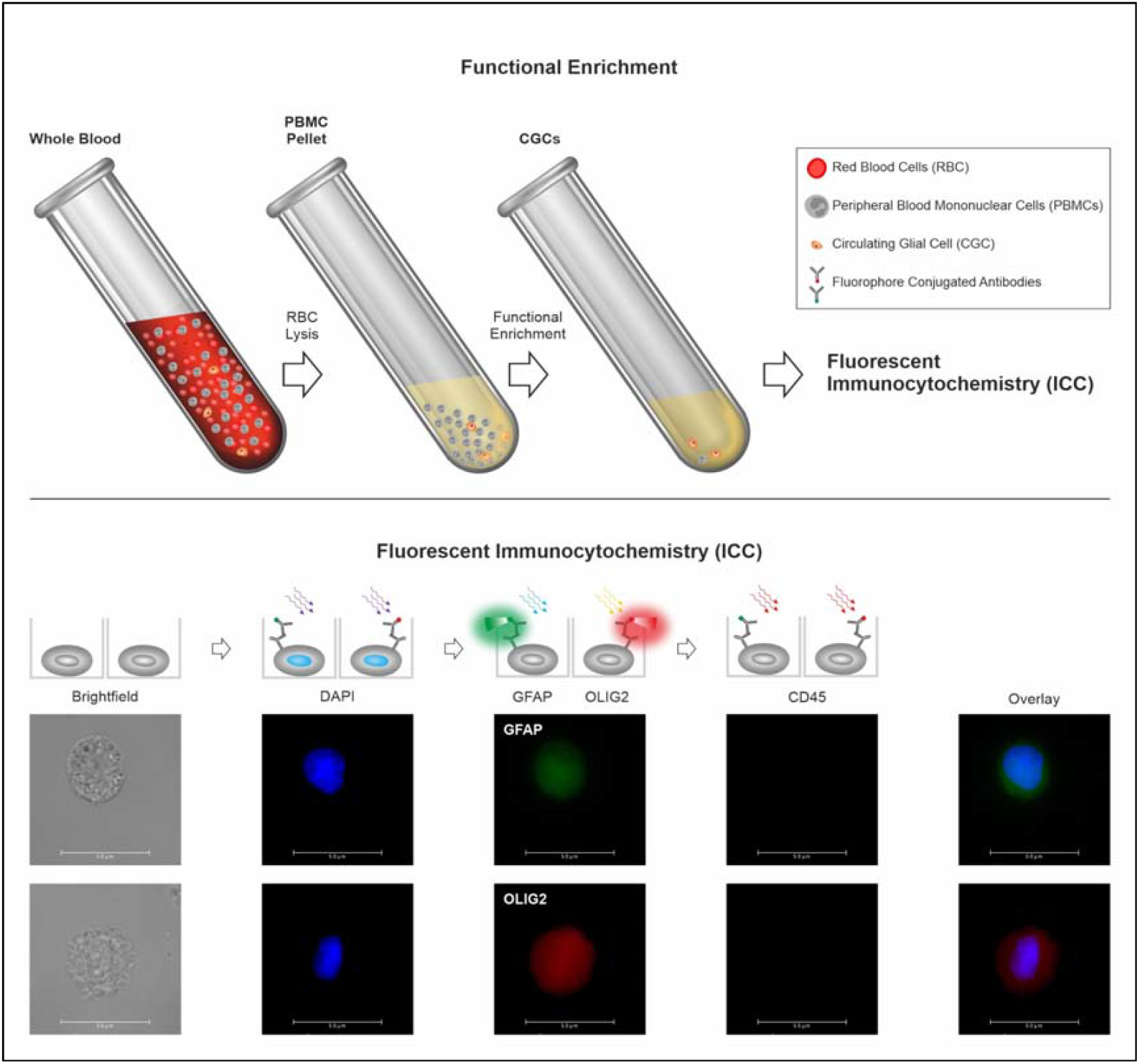
Schema of Test. Functional enrichment of CTCs is achieved using a proprietary CGC/CTC enrichment medium (CEM) that eliminates all non-malignant cells and permits tumor derived malignant cells to survive. Subsequently, the multiplexed immunocytochemistry (ICC) evaluates and identifies CGCs based on positivity of the indicated markers.

### Method Development and Optimization

#### Detection Thresholds

Approxiately 100 cells of each of the following types were seeded into imaging compatible 96 well plates and immunostained to determine the expression (FI) of GFAP, OLIG2 and CD45. Sample types included U87MG, MOLT-3 cells, CGCs, malignant (glial) tumor derived cells (M-TDCs) and non-malignant brain tumor derived cells (B-TDCs).

#### Marker Specificity

The specificity of GFAP and OLIG2 to glial malignancies was evaluating by determinin their expression (FI) in CTCs from various (non-CNS) primary malignancies which were seened into imaging compatible 96 well plates and immunostained for these markers.

#### Marker Expression in Glial Malignancies

The expression (FI) of GFAP and OLIG2 were evaluated in CGCs from subsets of patients stratified by gender and age-group (n = 34) as well as histological subtype and grade (n = 63).

### Analytical Validation

Analytical validation established the performance characteristics of the test with U87MG (glioblastoma reference cell line) cells. U87MG has been previously verified for high expression of GFAP and OLIG2 as well as absence of CD45 expression. Reference samples were generated by spiking measured amounts of U87MG cells into healthy donor blood (HDB) samples, processed for enrichment of apoptosis reluctant cells using the CEM and the enriched cells were used for ICC profiling to determine the recovery of GFAP and OLIG2 positive cells.

#### Analyte Stability and Recovery

To determine the Analyte Stability, 36 × 5 mL aliquots of healthy donor blood were each spiked with ∼15 U87MG cells. Of these 36 spiked samples, 6 samples each were either processed immediately or after 24 h, 48 h, 72 h, 96 h and 120 h storage at 2°C – 8°C respectively. Of the 6 aliquots evaluated at each time point, 3 aliquots each were used to determine recovery of GFAP+ cells and OLIG2+ cells respectively. Additionally, 30 mL blood was collected from a known case of CNS malignancy and split into 6 aliquots of 5 mL each; one sample was processed immediately (0 h), and the others were processed after 24 h, 48 h, 72 h, 96 h and 120 h storage at 2°C - 8°C respectively. Recovery at 0 h was normalized as 100% and recoveries at 24 h, 48 h, 72 h, 96 h and 120 h were determined relative to the 0 h recovery.

#### Linearity

U87MG cells were spiked into 176 × 5 mL aliquots of healthy donor blood samples, stored for 48 h at 2°C – 8°C and then processed for recovery. The 176 aliquots comprised 2 sets of 88 aliquots (11 spikes × 8 replicates). The study also included 16 × 5 mL aliquots (2 sets × 8 replicates) of healthy donor blood samples which were not spiked. Each set was assigned for detection of one of the two marker positive cell types (GFAP+ / OLIG2+). Samples were stored for 48 h at 2°C – 8°C. The Linearity Interval was evaluated as per the approach described in Clinical and Labpratory Standards Institute (CLSI) approved guideline EP06 (38). In addition, the linear response characteristics were also evaluated by Linear Regression to determine the coefficient of correlation (R^2^).

#### Limits of Detection, Quantitation and Blank

The Limit of Blank (LoB) was determined from the 8 × 5 mL unspiked healthy donor blood samples per marker in the Linearity study. The Limit of Detection (LoD) of each marker was determined from a subset of the Linearity Study which included 24 × 5 mL samples spiked with 1, 3 or 5 U87MG cells (8 of each). The LoQ was determined from a subset of 32 × 5 mL samples from the Linearity Study which were spiked with 1, 3, 5, or 10 U87MG cells (8 of each) per marker. The LoB, LoD and LoQ were determined as per the approach described in CLSI approved guideline EP17-A2 (39).

#### Sensitivity, Specificity and Accuracy

Sensitivity was determined from a subset of the Linearity Study samples which included 40 × 5 mL samples spiked with 5, 10, 20, 40 and 80 U87MG cells (8 of each) per marker type. Specificity was determined from the 8 × 5 mL unspiked healthy donor blood samples (per marker type) in the Linearity study. Accuracy was determined based on total true positive and true negative samples detected out of the total samples per marker type.

#### Precision

Precision of the test was evaluated at 5 cells / 5 mL (sample positivity threshold) as well as at 15 cells / 5 mL (3 × threshold) using a 10 × 2 × 8 design which yielded a total of 160 observations on eplicate samples over 10 days, from 8 replicates of 2 users each. Contrived samples for the precision study were generated by User 1 by spiking 5 and 15 U87MG cells into separate 8 × 5 mL aliquots of healthy donor blood (HDB) per day for 10 days. Samples were processed by CEM treatment and ICC profiling to determine recovery of marker positive cells. User 2 repeated the above study at both spike densities on the same 10 days. Mean of observed recoveries were used to calculate Standard Deviation (SD) and Coefficient of Variation (CV, %) for repeatability and within-laboratory precision (since this was a single site study) as per the two-factor nested analysis of variance (ANOVA) approach described in CLSI approved guideline EP05-A3 (40).

#### Interfering Substances

The performance characteristics of the test were evaluated in presence of endogenous (pathology markers) and exogenous factors (non-anticancer drugs) as potential interfering agents. Pure (analytical grade) molecules for each of these agents were obtained from commercial vendors and stored under recommended conditions until use. All substances were reconstituted as per manufacturer’s instructions in appropriate solvents to prepare working stock solutions which were immediately used for spiking studies. The exogenous substances (drugs) were used at the reported medically relevant Peak Plasma Concentrations (C_Max_), while endogenous substances (serum parameters) were evaluated at concentrations that are considered clinically elevated. Blood from a healthy donor (120 mL) who was not under any medication (last 14 days) was procured from a blood bank and spiked with about 1200 U87MG cells (to achieve 10 cells / mL). The spiked sample was split into 24 × 5 mL aliquots; 23 aliquots were spiked with each of the above substances at the indicated concentrations and 1 aliquot was used as an unspiked control. Samples were processed by CEM treatment and ICC profiling to determine recovery of marker positive cells.

### Clinical Studies

The ability of the test to identify and differentiate GLI-M from NBT based on assessment of GFAP and OLIG2 was first ascertained and established in a cohort of 189 samples which included 145 known cases (recently diagnosed, therapy naïve) of GLI-M, and 44 known cases of NBT (Supplementary Table S7). All samples were assigned to Training and Test Sets in a 70%:30% ratio. The analysts were unblinded to the clinical status of samples in the Training Set to determine the concordance of marker expression (Decision Matrix). Subsequently, the analysts who remained blinded to the actual clinical status of samples in the Test Set, predicted the status of these samples based on the marker expression profiles and the Decision Matrix. The concordance of the prediction with the actual clinical status (which was subsequently revealed) was used to determine the performance characteristics of the test.

The ability of the test to identify and differentiate GLI-M from NBT and EPI-M based on assessment of GFAP, OLIG2 and PanCK was ascertained and established in a second cohort of 586 samples which included 500 healthy adults (no prior diagnosis of cancer nor any current symptoms or clinical features of cancer), 24 previously diagnosed and treated cases of EPI-M with brain metastases, 40 recently diagnosed therapy naïve cases of GLI-M and 22 known cases of NBT (Supplementary Table S11). After sample processing and ICC, the analysts predicted the status of these samples based on the marker expression profiles and the Decision Matrix. The concordance of the prediction with the actual clinical status (which was subsequently revealed) was used to determine the performance characteristics of the test.

The performance characteristics of the test were evaluated in a prospective multi-centre cohort of 68 individuals presenting with ICSOL on brain imaging suspected of GLI-M (Supplementary Table S14). After sample processing and ICC, the analysis predicted the status of these samples based on the marker expression profiles and the Decision Matrix. The concordance of the prediction with the actual clinical status (which was subsequently revealed) was used to determine the performance characteristics of the test.

The performance characteristics of the test were evaluated in a second prospective cohort of 31 individuals presenting with intra-axial ICSOL on brain imaging (Supplementary Table S18). After sample processing and ICC, the analysis predicted the status of these samples based on the marker expression profiles and the Decision Matrix. The concordance of the prediction with the actual clinical status (which was subsequently revealed) was used to determine the performance characteristics.

### Orthogonal Verification – Fluorescence in situ Hybridization (FISH)

FISH was performed on enriched malignant cells from pre-biopsy blood samples and matched tumor tissue from 44 individuals including 22 cases of GLI-M and 22 cases of NBT to determine concordance in detection of EGFR gene amplification. This cohort of samples was populated from the three clinical studies where matched blood and tumor tissue samples were available for each patient. FISH was performed according to manufacturer’s instructions using ZytoLight^®^ SPEC EGFR/CEN 7 Dual Color Probe (Zytovision Inc, Germany), which is a mixture of ZyOrange (Excitation: 547 nm; Emission: 572 nm) conjugated “CEN 7” probe specific for the alpha satellite centromeric region of Chromosome 7 (D7Z1) and ZyGreen (Excitation: 503 nm; Emission: 528 nm) conjugated “SPEC EGFR” probe specific for the chromosomal region 7p11.2* harboring the EGFR gene. The protocol for processing tumor tissue as well as malignant cells in suspension (CTCs / CGCs) was as specified in the manufacturer’s instructions for use of the kit. A-431 (vulval squamous cell carcinoma) cell line which has been reported to harbour EGFR amplification was used as positive control as were tumor samples where EGFR amplification was previously ascertained by next generation sequencing (NGS). Peripheral blood mononuclear cells (PBMCs) isolated from asymptomatic individuals with no prior history of cancer and no current symptoms suspected of cancer were used as negative controls. Processed samples were visualized by fluorescence microscopy (Axio Imager Z2, Carl Zeiss, Germany). Reference cell line A-431 was used for validation of EGFR FISH probes.

## Data Availability

All data produced in the present work are contained in the manuscript and its supplementary file.

## AUTHOR CONTRIBUTIONS

Kevin O’Neill: Conceptualization, Investigation, Methodology, Project administration, Resources, Writing – review & editing. Nelofer Syed: Conceptualization, Investigation, Methodology, Project administration, Resources, Writing – review & editing. Timothy Crook: Conceptualization, Investigation, Methodology, Project administration, Resources, Writing – review & editing. Sudhir Dubey: Conceptualization, Investigation, Writing – review & editing. Mahadev Potharaju: Conceptualization, Investigation, Writing – review & editing. Sewanti Limaye: Conceptualization, Investigation, Writing – review & editing. Anantbhushan Ranade: Conceptualization, Investigation, Writing – review & editing. Giulio Anichini: Conceptualization, Investigation, Project administration, Resources, Writing – review & editing. Vineet Datta: Conceptualization, Funding acquisition, Investigation, Methodology, Project administration, Resources, Software,Supervision, Validation, Visualization, Writing – original draft.

All authors have read the manuscript and agreed its contents, the author list and its order as well as the above author contribution statements.

## ACKNOWLEDGEMENTS

The authors acknowledge gratitude to the patients who participated in the studies as well as the families of these patients. The authors also acknowledge the contributions of the clinical staff at the participating sites and employees of the study sponsor for their contributions in managing various clinical, operational and laboratory aspects of the studies.

## DATA AVAILABILITY

De-identified data including patient (study participant) information as well as data generated from patient samples cannot be shared because patient consent has not been obtained for sharing of de-identified data with third parties (not directly / indirectly involved in the studies). Proprietary data belonging to the study sponsor can not be shared due to intellectual property considerations. Data arising from analysis of primary data and any additional analyzed data can be made available upon e-mail request sent to the corresponding author, after execution of confidentiality and non-disclosure agreement(s), and will be subject to restrictions on further use (reuse, reanalysis) or further dissemination in any form.

## CODE AVAILABILITY

No commercial, open source or custom codes have been used for data capture. Data was analyzed as per the statistical analyses described in the CLSI guidelines (which are cited in the manuscript) using MS Excel spreadsheets the MedCalc Statistical Calculator (https://www.medcalc.org/calc/diagnostic_test.php). No other (commercial, open source or custom) codes were used for data analysis.

## MATERIALS AVAILABILITY

Human samples described in this manuscript can not be made available to third parties since consent has not been obtained from study participants and volunteers to share these samples with third parties. Proprietary formulations of the study sponsor cannot be made available to third parties due to intellectual property considerations. Other biological materials described in the manuscript can be procured from commercial vendors.

## REFERENCES

1. Brain Tumor: Statistics. Cancer.Net. [cited 2022 Feb 19]. Available from: https://www.cancer.net/cancer-types/brain-tumor/statistics

2. Cancer Stat Facts: Brain and Other Nervous System Cancer [Internet]. Seer.cancer.gov. [cited 2022 Feb 19]. Available from: https://seer.cancer.gov/statfacts/html/prost.html

3. Miller KD, et al. Brain and other central nervous system tumor statistics, 2021. CA Cancer J Clin. 2021 Sep;71(5):381–406. Doi: 10.3322/caac.21693.

4. Malone H, Yang J, Hershman DL, Wright JD, Bruce JN, Neugut AI. Complications Following Stereotactic Needle Biopsy of Intracranial Tumors. World Neurosurg. 2015 Oct;84(4):1084–9.

5. Koszewski W, Kroh H, Kunert P. [Difficulties in stereotactic biopsies of brain tumors]. Neurol Neurochir Pol. 2002;36(3):481–8.

6. Ramkissoon LA, Pegram W, Haberberger J, Danziger N, Lesser G, Strowd R, et al. Genomic Profiling of Circulating Tumor DNA From Cerebrospinal Fluid to Guide Clinical Decision Making for Patients With Primary and Metastatic Brain Tumors. Front Neurol. 2020;11:544680.

7. Nassiri F, Chakravarthy A, Feng S, Shen SY, Nejad R, Zuccato JA, et al. Detection and discrimination of intracranial tumors using plasma cell-free DNA methylomes. Nat Med. 2020 Jul;26(7):1044–7.

8. Ebrahimkhani S, Vafaee F, Hallal S, Wei H, Lee MYT, Young PE, et al. Deep sequencing of circulating exosomal microRNA allows non-invasive glioblastoma diagnosis. NPJ Precis Oncol. 2018;2:28.

9. Pickles JC, Fairchild AR, Stone TJ, Brownlee L, Merve A, Yasin SA, et al. DNA methylation-based profiling for paediatric CNS tumour diagnosis and treatment: a population-based study. Lancet Child Adolesc Heal. 2020 Feb;4(2):121–30.

10. Macarthur KM, Kao GD, Chandrasekaran S, Alonso-Basanta M, Chapman C, Lustig RA, et al. Detection of brain tumor cells in the peripheral blood by a telomerase promoter-based assay. Cancer Res. 2014 Apr;74(8):2152–9.

11. Müller C, Holtschmidt J, Auer M, Heitzer E, Lamszus K, Schulte A, et al. Hematogenous dissemination of glioblastoma multiforme. Sci Transl Med. 2014 Jul;6(247):247ra101.

12. Kan LK, Drummond K, Hunn M, Williams D, O’Brien TJ, Monif M. Potential biomarkers and challenges in glioma diagnosis, therapy and prognosis. BMJ Neurol open. 2020;2(2):e000069.

13. Akolkar D, Patil D, Crook T, Limaye S, Page R, Datta V, et al. Circulating ensembles of tumor-associated cells: A redoubtable new systemic hallmark of cancer. Int J cancer. 2020 Jun;146(12):3485–94.

14. Ranade A, Bhatt A, Page R, Limaye S, Crook T, Akolkar D, et al. Hallmark Circulating Tumor-Associated Cell Clusters Signify 230 Times Higher One-Year Cancer Risk. Cancer Prev Res (Phila). 2021 Jan;14(1):11–6.

15. Gaya A, Crook T, Plowman N, Ranade A, Limaye S, Bhatt A, et al. Evaluation of circulating tumor cell clusters for pan-cancer noninvasive diagnostic triaging. Cancer Cytopathol. 2021 Mar;129(3):226–38.

16. Hamilton JD, Rapp M, Schneiderhan T, Sabel M, Hayman A, Scherer A, et al. Glioblastoma multiforme metastasis outside the CNS: three case reports and possible mechanisms of escape. J Clin Oncol Off J Am Soc Clin Oncol. 2014 Aug;32(22):e80–4.

17. Pasquier B, Pasquier D, N’Golet A, Panh MH, Couderc P. Extraneural metastases of astrocytomas and glioblastomas: clinicopathological study of two cases and review of literature. Cancer. (1980) 45:112–25. 10.1002/1097-0142(19800101)45:1<112::AID-CNCR2820450121>3.0.CO;2-9

18. Chang H, Ding Y, Wang P, Wang Q, Lin Y, Li B. Cutaneous metastases of the glioma. J Craniofac Surg. (2018) 29:e94–e6. 10.1097/SCS.0000000000004204

19. Perez-Bovet J, Rimbau-Munoz J. Glioblastoma multiforme metastases to the masticator muscles and the scalp. J Clin Neurosci. (2018) 53:237–9. 10.1016/j.jocn.2018.04.021

20. Rosen J, Blau T, Grau SJ, Barbe MT, Fink GR, Galldiks N. Extracranial metastases of a cerebral glioblastoma: a case report and review of the literature. Case Rep Oncol. (2018) 11:591–600. 10.1159/000492111

21. Eibl RH, Schneemann M. Liquid Biopsy and Primary Brain Tumors. Cancers (Basel). 2021 Oct;13(21).

22. Zhang H, Yuan F, Qi Y, Liu B, Chen Q. Circulating Tumor Cells for Glioma. Front Oncol. 2021 Mar 10;11:607150. Doi: 10.3389/fonc.2021.607150.

23. Bang-Christensen SR, Pedersen RS, Pereira MA, Clausen TM, Løppke C, Sand NT, et al. Capture and Detection of Circulating Glioma Cells Using the Recombinant VAR2CSA Malaria Protein. Cells. 2019 Aug;8(9).

24. Sullivan JP, Nahed B V, Madden MW, Oliveira SM, Springer S, Bhere D, et al. Brain tumor cells in circulation are enriched for mesenchymal gene expression. Cancer Discov. 2014 Nov;4(11):1299–309.

25. Gao F, Cui Y, Jiang H, Sui D, Wang Y, Jiang Z, et al. Circulating tumor cell is a common property of brain glioma and promotes the monitoring system. Oncotarget. 2016 Nov;7(44):71330–40.

26. Krol I, Castro-Giner F, Maurer M, Gkountela S, Szczerba BM, Scherrer R, et al. Detection of circulating tumour cell clusters in human glioblastoma. Br J Cancer. 2018 Aug;119(4):487–91.

27. Crook T, Leonard R, Mokbel K, Thompson A, Michell M, Page R, Vaid A, Mehrotra R, Ranade A, Limaye S, Patil D, Akolkar D, Datta V, Fulmali P, Apurwa S, Schuster S, Srinivasan A, Datar R. Accurate Screening for Early-Stage Breast Cancer by Detection and Profiling of Circulating Tumor Cells. Cancers (Basel). 2022 Jul 9;14(14):3341. Doi: 10.3390/cancers14143341.

28. Limaye S, Chowdhury S, Rohatgi N, Ranade A, Syed N, Riedemann J, Patil D, Akolkar D, Datta V, Patel S, Chougule R, Shejwalkar P, Bendale K, Apurwa S, Schuster S, John J, Srinivasan S, Datar R. Accurate Prostate Cancer Detection based on Enrichment and Characterization of Prostate Cancer Specific Circulating Tumor Cells. Cancer Medicine, 2023. Accepted. In Press.

29. Fazeny-Dörner, B., et al. (2003). Survival and prognostic factors of patients with unresectable glioblastoma multiforme. Anti-Cancer Drugs, 14(4), 305–312. doi:10.1097/00001813-200304000-00008

30. Air EL, et al. Management strategies after nondiagnostic results with frameless stereotactic needle biopsy: Retrospective review of 28 patients. Surg Neurol Int. 2012;3(Suppl 4):S315–S319. doi:10.4103/2152-7806.103026

31. Woodworth GF, et al. Frameless image-guided stereotactic brain biopsy procedure: diagnostic yield, surgical morbidity, and comparison with the frame-based technique. J Neurosurg. 2006 Feb;104(2):233–7. doi: 10.3171/jns.2006.104.2.233

32. Khatab, S., et al. (2014). Frameless image-guided stereotactic brain biopsies: emphasis on diagnostic yield. Acta Neurochirurgica, 156(8), 1441–1450. doi:10.1007/s00701-014-2145-2.

33. Füreder LM, Widhalm G, Gatterbauer B, Dieckmann K, Hainfellner JA, Bartsch R, Zielinski CC, Preusser M, Berghoff AS. Brain metastases as first manifestation of advanced cancer: exploratory analysis of 459 patients at a tertiary care center. Clin Exp Metastasis. 2018 Dec;35(8):727–738. doi: 10.1007/s10585-018-9947-1.

34. Louis DN, Perry A, Wesseling P, Brat DJ, Cree IA, Figarella-Branger D, Hawkins C, Ng HK, Pfister SM, Reifenberger G, Soffietti R, von Deimling A, Ellison DW et al. The 2021 WHO Classification of Tumors of the Central Nervous System: a summary. Neuro Oncol. 2021 Aug 2;23(8):1231–1251. doi: 10.1093/neuonc/noab106.

35. Kolostova K, Pospisilova E, Pavlickova V, Bartos R, Sames M, Pawlak I, Bobek V. Next generation sequencing of glioblastoma circulating tumor cells: non-invasive solution for disease monitoring. Am J Transl Res. 2021 May 15;13(5):4489–4499.

36. Von Elm E, at al. The Strengthening the Reporting of Observational Studies in Epidemiology (STROBE) Statement: guidelines for reporting observational studies. Int J Surg 2014;12:1495–9. DOI: 10.1016/j.ijsu.2014.07.013.

37. Crook T, Gaya A, Page R, Limaye S, Ranade A, Bhatt A, et al. Clinical utility of circulating tumor-associated cells to predict and monitor chemo-response in solid tumors. Cancer Chemother Pharmacol. 2021 Feb;87(2):197–205.

38. CLSI. Evaluation of Linearity of Quantitative measurement procedures. 2nd ed. CLSI Guideline EP06. Clinical and Labpratory Standards Institute; 2020.

39. CLSI. Evaluation of Detection Capability for Clinical Laboratory Measurement Procedures.; Approved Guideline. 2nd ed. CLSI document EP17-A2. Clinical and Labpratory Standards Institute; 2012

40. CLSI. Evaluation of Detection Capability for Clinical Laboratory Measurement Procedures.; Approved Guideline. 2nd ed. CLSI document EP17-A2. Clinical and Labpratory Standards Institute; 2012.

