## Supplementary Figures and Tables for "Profiling of circulating glial cells allows accurate blood-based diagnosis of glial malignancies"

**Supplementary Figure S1. Detection Thresholds**. The expression level (range of fluorescent intensity, FI) of each marker was evaluated on MOLT-3, SKBR3, U87MG, CGCs, malignant glial tumor derived cells (M-TDC) and non-malignant brain tumor cells (B-TDC). The box and whisker plots indicate the range, intra-(25^th^-75^th^)-quartile range and median (horizontal line within box). The FI range observed in this study formed the basis for the detection threshold for each marker on CGCs.

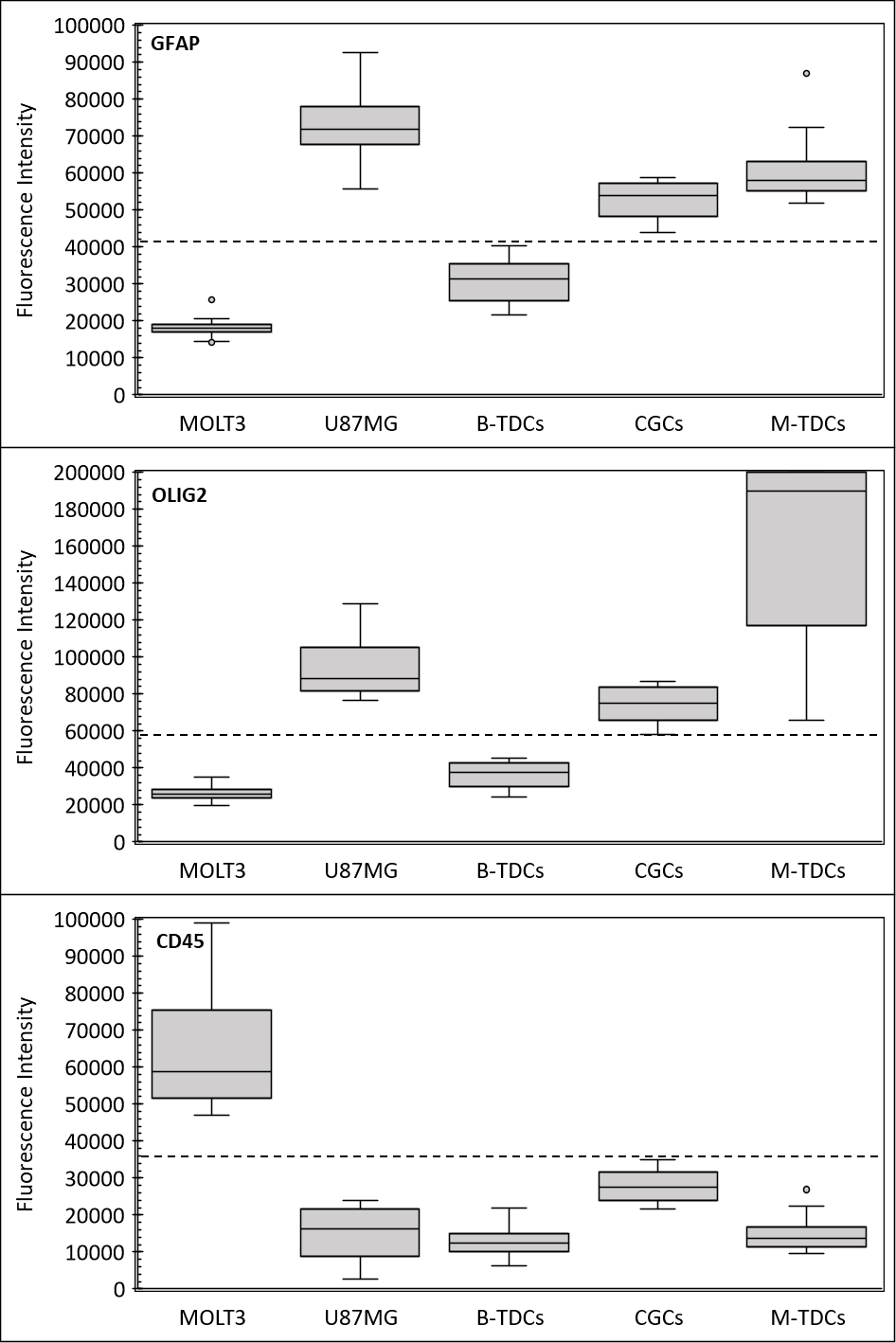

**Supplementary Figure S2. GFAP and OLIG2 Expression in CTCs from other (non-CNS) malignancies.** CTCs from various (non-CNS) cancer types were evaluated for and found to be negative for GFAP and OLIG2 expression (FI < 45,000).

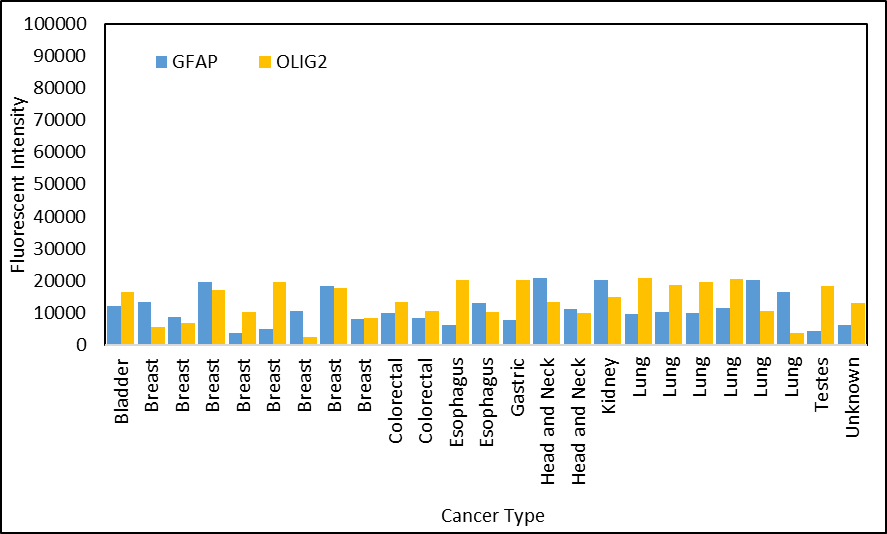

**Supplementary Figure S3. Gender and Age-group and Marker Expression on CGCs.** CGCs from individuals with GLI-M were evaluated for differences in expression levels (fluorescence intensity, FI) of GFAP and OLIG2 based on age group (less than 20 years, 20 – 40 years, 40 – 60 years and 60 – 80 years) or gender (F: female and M: male). Neither age nor gender was associated with any loss of sensitivity for any marker. The box and whisker plots indicate the range, intra-(25^th^-75^th^)-quartile range, median (horizontal line within box) and mean (x within box).

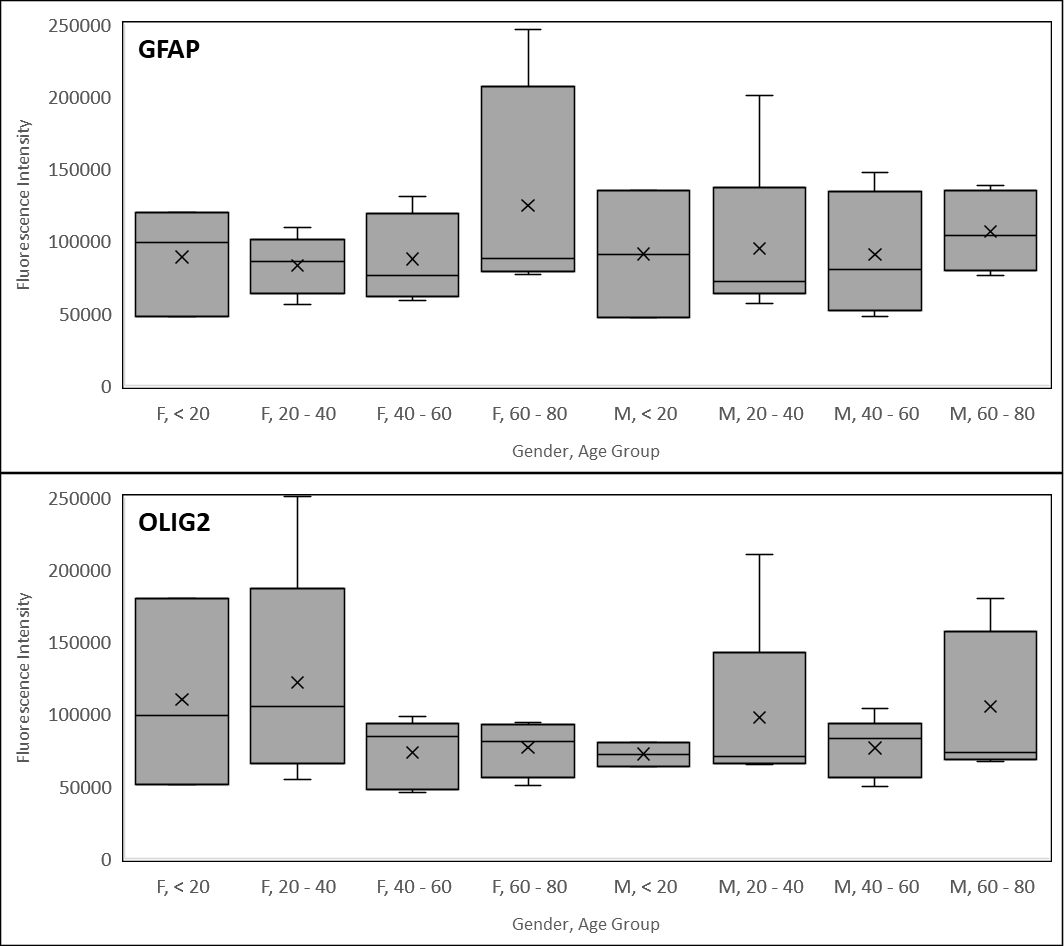

**Supplementary Figure S4. Subtype and Marker Expression on CGCs.** CGCs from individuals with various subtypes and grade of GLI-M were evaluated for differences in expression levels (fluorescence intensity, FI) of GFAP and OLIG2 based on subtype (Ast: Astrocytoma; Epe: Enendymoma; GBM: Glioblastoma multiforme; Gli: Glioma Not Otherwise Specified; Oli: Oligodendroglioma) or grade (II, III, IV). Neither subype nor grade of tumor was associated with any loss of sensitivity for any marker. Ependymomas are negative for OLIG2 and their detection is based on GFAP positivity alone. The box and whisker plots indicate the range, intra-(25^th^-75^th^)-quartile range, median (horizontal line within box) and mean (x within box).

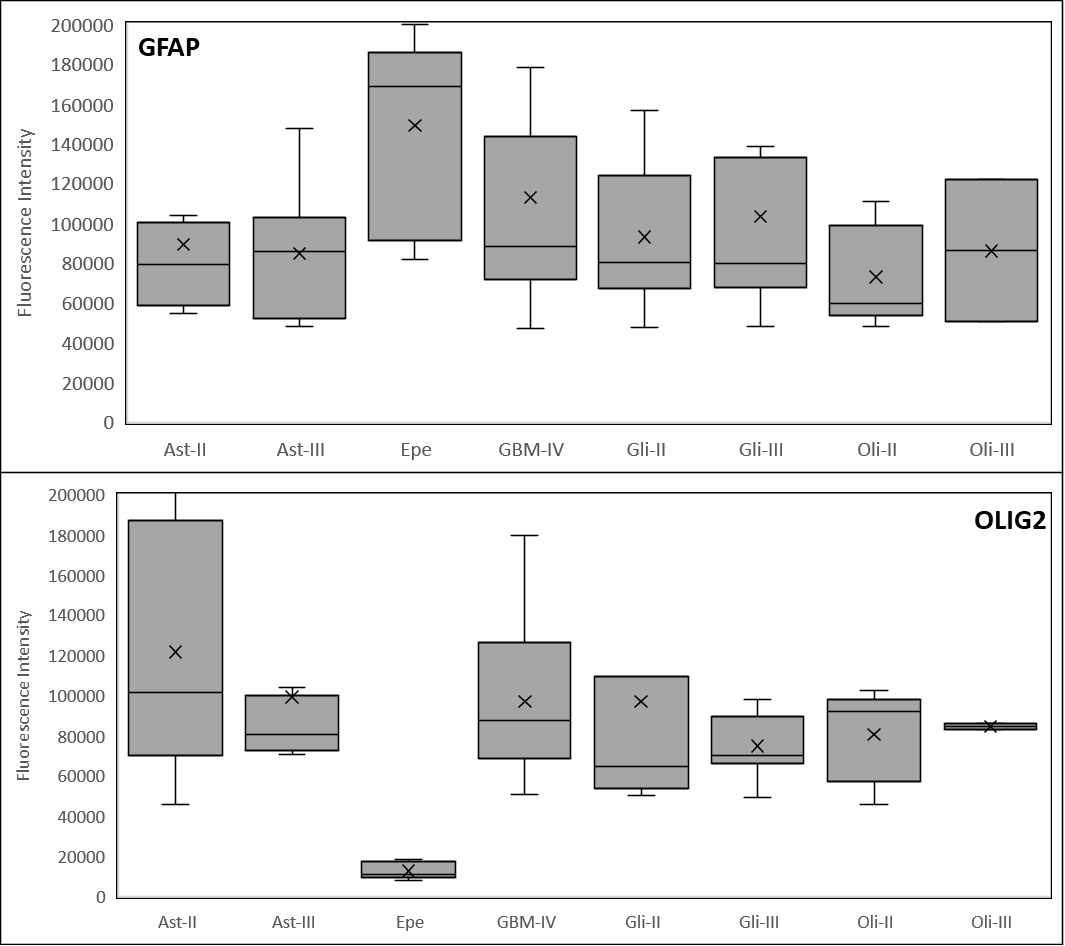

**Supplementary Figure S5: Analytical Validation: Linearity.** The Test exhibited significant linearity with R^2^ ≥0.99.

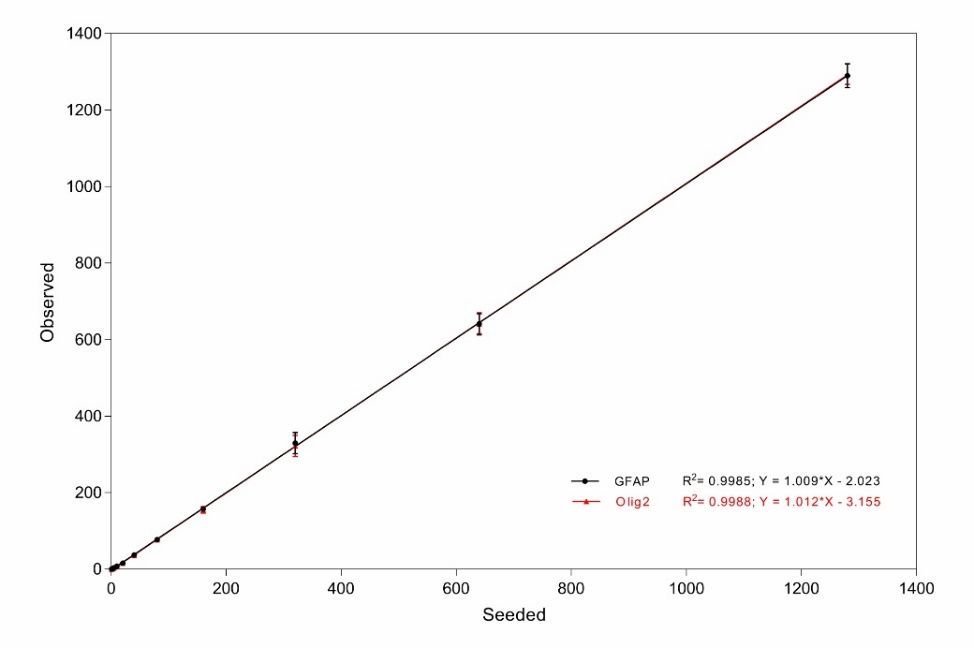

**SUPPLEMENTARY TABLES**

**Supplementary Table S1. Analytical Validation: Analyte Stability (Spiked Samples)**. U87MG cells were spiked into healthy donor blood samples and the recovery of spiked cells was evaluated for up to 120 hours.

| **Time (h)** | **Spiked** | **Recovery of GFAP+ Cells** | | **Recovery of OLIG2+ Cells** | |
| --- | --- | --- | --- | --- | --- |
| **Spiked Samples** | | **Mean (%)** | **Range** | **Mean (%)** | **Range** |
| 0 | 15 | 13 (86.7%) | (12-13) | 13 (86.7%) | (12-14) |
| 24 | 15 | 12 (80.0%) | (12-13) | 12 (80.0%) | (12-13) |
| 48 | 15 | 12 (80.0%) | (12-12) | 12 (80.0%) | (11-13) |
| 72 | 15 | 12 (80.0%) | (11-13) | 12 (80.0%) | (11-12) |
| 96 | 15 | 9 (60.0%) | (8-10) | 8 (53.3%) | (7-9) |
| 120 | 15 | 8 (53.3%) | (7-9) | 9 (60.0%) | (8-11) |
| **Clinical Samples** | | **Recovery** | **Relative %** | **Recovery** | **Relative %** |
| 0 | - | 7 | (100%) | 9 | (100%) |
| 24 | - | 6 | 85.7% | 8 | 88.9% |
| 48 | - | 6 | 85.7% | 8 | 88.9% |
| 72 | - | 6 | 85.7% | 7 | 77.8% |
| 96 | - | 4 | 57.1% | 4 | 44.4% |
| 120 | - | 2 | 28.6% | 1 | 11.1% |

**Supplementary Table S2: Analytical Validation: Linearity.** The Tabulated values indicaqte the recovery and range of recovery in the linearity study samples.

| **Spiked Cells** | **Recovery of GFAP+ Cells** | | **Recovery of OLIG2+ Cells** | |
| --- | --- | --- | --- | --- |
|  | **Mean (%)** | **Range** | **Mean (%)** | **Range** |
| 0 | 0 | - | 0 | - |
| 1 | 0.4 (37.5%) | 0 – 1 | 0.4 (37.5%) | 0 – 1 |
| 3 | 1.4 (45.8%) | 1 – 2 | 2.3 (75.0%) | 2 - 3 |
| 5 | 3.9 (77.5%) | 3 – 4 | 3.9 (77.5%) | 3 – 4 |
| 10 | 7.5 (75.0%) | 6 – 8 | 7.6 (76.3%) | 7 - 8 |
| 20 | 15.8 (78.8%) | 15 – 17 | 15.5 (77.5%) | 14 - 18 |
| 40 | 37.1 (92.8%) | 34 – 40 | 35.9 (89.7%) | 32 - 43 |
| 80 | 77.1 (96.4%) | 69 – 87 | 77.0 (96.3%) | 69 - 84 |
| 160 | 157.4 (98.4%) | 145 – 166 | 154.8 (96.7%) | 142 - 168 |
| 320 | 329.1 (102.9%) | 285 – 381 | 322.1 (100.7%) | 278 - 355 |
| 640 | 641.0 (100.2%) | 609 – 697 | 641.0 (100.2%) | 600 - 672 |
| 1280 | 1289.4 (100.7%) | 1245 - 1330 | 1293.4 (101.0%) | 1260 - 1330 |

**Supplementary Table S3. Analytical Validation: Sensitivity, Specificity, Accuracy.** U87MG cells were spiked into healthy donor blood samples at various seed densities (8 replicates each) and their recoveries evaluated to determine sensitivity (proportion of samples with true positive findings). Unspiked healthy donor blood samples were evaluated for false positives to determine specificity (proportion of samples with true negative findings). Accuracy was determined from sensitivity and specificity.

| **Sample Type** | **Detected Cells: Mean (Range)** | **Number of Samples** | | | **Performance** | |
| --- | --- | --- | --- | --- | --- | --- |
|  |  | **Total** | **Positive*** | **Negative** | **Sensitivity** | **Specificity** |
| **GFAP+, CD45-** | | | | | | |
| **Unspiked** | - | **8** | **-** | **8** | **-** | **100%** |
| **Spiked**  *5*  *10*  *20*  *40*  *80* | *4 (3-5)*  *8 (6-10)*  *16 (11-22)*  *36 (29-44)*  *77 (73-85)* | **40**  *8*  *8*  *8*  *8*  *8* | **37**  *5*  *8*  *8*  *8*  *8* | **3**  *3*  *-*  *-*  *-*  *-* | **92.5%**  *62.5%*  *100.0%*  *100.0%*  *100.0%*  *100.0%* | - |
| **OLIG2+, CD45-** | | | | | | |
| **Unspiked** | - | **8** | **-** | **8** | **-** | **100%** |
| **Spiked**  *5*  *10*  *20*  *40*  *80* | *4 (2-6)*  *8 (6-11)*  *17 (11-22)*  *36 (32-44)*  *73 (65-82)* | **40**  *8*  *8*  *8*  *8*  *8* | **38**  *6*  *8*  *8*  *8*  *8* | **2**  *2*  *-*  *-*  *-*  *-* | **95.0%**  *75.0%*  *100.0%*  *100.0%*  *100.0%*  *100.0%* | - |
| **Positive samples are those where ≥4 marker positive cells were detected* | | | | | | |

**Supplementary Table S4. Analytical Validation: Precision.** Summary of findings of the single site precision study based on the 10 × 2 × 8 design for determination of repeatability and within laboratory precision at 5 cells / 5 mL (detection threshold) and 15 cells / 5 mL (positive sample at 3× detection threshold) by two-factor nested ANOVA. SD: standard deviation; CV: coefficient of variation (%)

| **Spike Density** | **Mean of Observations** | **Repeatability** | | **Within Laboratory Precision** | |
| --- | --- | --- | --- | --- | --- |
|  |  | **SD** | **%CV**  **(95% CI)** | **SD** | **%CV**  **(95% CI)** |
| 5 | 5.9 | 0.80 | 13.7%  (11.9% - 16.2%) | 1.38 | 23.5%  (17.4% - 37.4%) |
| 15 | 15.8 | 1.59 | 10.0%  (8.7% - 11.9%) | 2.17 | 13.7%  (10.4% - 19.9%) |
| **values indicate number of cells / 5 mL* | | | | | |

**Supplementary Table S5. Analytical Validation: Impact of Potentially Interfering Substances.** The Test was not prone to interference from endogenous agents (deranged serum parameters) and exogenous agents (common non-anticancer drugs).

| **Agent** | **Concentration Used** | **Detected Cells / mL** | |
| --- | --- | --- | --- |
|  |  | **GFAP+** | **OLIG2+** |
| Levothyroxine | 140 ng / mL | 10 | 9 |
| Lisinopril | 58 ng / mL | 8 | 8 |
| Atorvastatin | 30 ng / mL | 9 | 10 |
| Metformin | 5 μg / mL | 9 | 9 |
| Amlodipine | 5 ng / mL | 9 | 10 |
| Metoprolol | 50 ng / mL | 10 | 10 |
| Omeprazole | 660 ng / mL | 9 | 9 |
| Albuterol | 4.2 ng / mL | 9 | 8 |
| Ranitidine | 450 ng / mL | 8 | 9 |
| Azithromycin | 500 ng / mL | 10 | 8 |
| Paracetamol | 9.9 μg / mL | 9 | 8 |
| Aspirin | 3 μg / mL | 9 | 8 |
| Loperamide | 3.4 ng / mL | 10 | 9 |
| Dextromethorphan | 2.9 ng / mL | 8 | 10 |
| Ulipristal acetate | 170 ng / mL | 9 | 9 |
| Cortisone | 2700 nmol/l | 8 | 10 |
| Sildenafil | 440 ng/ml | 10 | 10 |
| Bilirubin | 20 μg / mL | 9 | 9 |
| Cholesterol | 3 mg / mL | 8 | 8 |
| Glucose | 3 mg / mL | 9 | 10 |
| Haemoglobin | 200 mg / mL | 9 | 9 |
| Uric Acid | 150 μg / mL | 9 | 10 |
| EDTA | 20 mg/mL | 9 | 10 |
| Control | - | 10 | 10 |

**Supplementary Table S6. Inclusion-Exclusion Criteria for Clinical Study-1.**

|  | **Inclusion Criteria** | **Exclusion Criteria** |
| --- | --- | --- |
| **GLI-M** | - Recently diagnosed cases, - Histopathological diagnosis available, - No prior radiation or systemic anticancer treatments, - Provision of informed consent, - Willing to provide blood sample, | - Inability to meet all the inclusion criteria, - Non glial CNS malignancies, - Non CNS malignancies with brain metastases, - Synchronous or metachronous malignancies, |
| **NBT** | - Recently diagnosed cases, - Histopathological diagnosis available, - Provision of informed consent, - Willing to provide blood sample, | - Inability to meet all the inclusion criteria, - Prior diagnosis of any malignancy, - Suspicion of non CNS malignancy |

**Supplementary Table S7. Demographics of the Clinical Study-1 Cohort.**

Samples from this cohort were divided into Training and Test Sets and then evaluated to determine the ability of the test to differentiate GLI-M samples from NBT samples.

|  | **Training Set** | **Validation Set** | **Total** |
| --- | --- | --- | --- |
| **GLI-M** | | | |
| **Age**  Median (Range) | 46 (4 - 82) | 50 (2 - 83) | 46 (2 - 83) |
| **Gender**  Male  Female | 60  41 | 28  16 | 88  57 |
| **Subtype**  *Astrocytoma, Gr II*  *Astrocytoma, Gr III*  *Ependymoma, Gr II*  *Ependymoma, Gr III*  *Glioblastoma, Gr IV*  *Glioma, mixed, Gr II*  *Glioma, NOS, Gr II*  *Glioma, NOS, Gr III*  *Oligodendroglioma, Gr II*  *Oligodendroglioma, Gr III*  ***TOTAL*** | *20*  *11*  *5*  *1*  *44*  *1*  *10*  *5*  *3*  *1*  **101** | *9*  *5*  *1*  *-*  *18*  *1*  *4*  *3*  *2*  *1*  **44** | *29*  *16*  *6*  *1*  *62*  *2*  *14*  *8*  *5*  *2*  **145** |
| **NBT** | | | |
| **Age**  Median (Range) | 46 (20 - 62) | 49 (21 - 67) | 46 (20 - 67) |
| **Gender**  Male  Female | 11  20 | 6  7 | 17  27 |
| **Subtype**  *Arachnoid cyst*  *Choroid Plexus Papilloma*  *CP Angle Epidermoid cyst*  *CP Angle Schwannoma*  *Craniopharyngioma*  *Hemangioblastoma*  *Meningiomas (Gr I)*  *Pituitary Adenoma*  *Trigeminal Schwannoma*  *Tuberculoma*  *Ventricular Colloid Cyst*  *Ventricular Epidermoid Cyst*  ***TOTAL*** | *1*  *-*  *-*  *5*  *1*  *1*  *19*  *2*  *-*  *1*  *1*  *-*  **31** | *-*  *1*  *1*  *1*  *1*  *-*  *6*  *1*  *1*  *-*  *-*  *1*  **13** | *1*  *1*  *1*  *6*  *2*  *1*  *25*  *3*  *1*  *1*  *1*  *1*  **44** |

**Supplementary Table S8. Clinical Study-1 Observations.** The table provides the summary of observations (status of CGCs in each type of sample) in the Training and Test Sets of Clinical Study-1.

| **Sample Type** | **Samples** | **Positive** | **Negative** |
| --- | --- | --- | --- |
| **Training Set** | | | |
| **NBT**  *Arachnoid cyst*  *Choroid Plexus Papilloma*  *CP Angle Schwannoma*  *Craniopharyngioma*  *Hemangioblastoma*  *Meningioma (Gr I)*  *Pituitary Adenoma*  *Tuberculoma*  *Ventricular Colloid Cyst*  *Ventricular Epidermoid Cyst* | **31**  *1*  *1*  *5*  *1*  *1*  *17*  *2*  *1*  *1*  *1* | **1 (3.2%)**  1  -  -  -  -  -  -  -  - | **30 (96.8%)**  -  *1*  *5*  *1*  *1*  *17*  *2*  *1*  *1*  *1* |
| **GLI-M**  *Astrocytoma, Gr II*  *Astrocytoma, Gr III*  *Ependymoma, Gr II*  *Ependymoma, Gr III*  *Glioblastoma, Gr IV*  *Glioma, mixed, Gr II*  *Glioma, NOS, Gr II*  *Glioma, NOS, Gr III*  *Oligodendroglioma, Gr II*  *Oligodendroglioma, Gr III* | **101**  *20*  *11*  *5*  *1*  *44*  *1*  *10*  *5*  *3*  *1* | **100 (99%)**  *20*  *11*  *4*  *1*  *44*  *1*  *10*  *5*  *3*  *1* | **1 (1%)**  -  -  1  -  -  -  -  -  -  - |
| **Test Set** | | | |
| **NBT**  *CP Angle Epidermoid Cyst*  *CP Angle Schwannoma*  *Craniopharyngioma*  *Meningioma*  *Pituitary Adenoma*  *Trigeminal Schwannoma* | **13**  *1*  *1*  *1*  *8*  *1*  *1* | **-**  -  -  -  -  -  - | **13 (100.0%)**  *1*  *1*  *1*  *8*  *1*  *1* |
| **GLI-M**  *Astrocytoma, Gr II*  *Astrocytoma, Gr III*  *Ependymoma, Gr II*  *Glioblastoma, Gr IV*  *Glioma, mixed, Gr II*  *Glioma, NOS, Gr II*  *Glioma, NOS, Gr III*  *Oligodendroglioma, Gr II*  *Oligodendroglioma, Gr III* | **44**  *9*  *5*  *1*  *18*  *1*  *4*  *3*  *2*  *1* | **44 (100%)**  *9*  *5*  *1*  *18*  *1*  *4*  *3*  *2*  *1* | **-**  -  -  -  -  -  -  -  -  - |

**Supplementary Table S9. Clinical Study-1 Performance.** The table summarizes the performance characteristics of the test in Clinical Study-1.

|  | **Number** | **Post unblinding** | |
| --- | --- | --- | --- |
|  |  | **NBT** | **GLI-M** |
| **Samples** | 57 | 13 | 44 |
| **Negative**  **(CGC-)** | 13 | 13 (100.0%) | - |
| **Positive**  **(CGC+)** | 44 | - | 44 (100.0%) |
| **Performance Characteristics** | | | |
| **Sensitivity** | - | 100.00%  (95%CI: 91.96% - 100.0%) | |
| **Specificity** | - | 100.00%  (95%CI: 75.29% - 100.0%) | |
| **Accuracy** | - | 100.00%  (95%CI: 93.73% - 100.0%) | |

**Supplementary Table S10. Inclusion-Exclusion Criteria for Clinical Study-2.**

|  | **Inclusion Criteria** | **Exclusion Criteria** |
| --- | --- | --- |
| **GLI-M** | - Histopathologically confirmed diagnosis, - Provision of informed consent, - Willing to provide blood sample, | - Inability to meet all the inclusion criteria, - Non glial CNS malignancies, - Non CNS malignancies with brain metastases, - Synchronous or metachronous malignancies, |
| **NBT** | - Histopathologically confirmed diagnosis, - Provision of informed consent, - Willing to provide blood sample, | - Inability to meet all the inclusion criteria, - Prior diagnosis of any malignancy, |
| **EPI-M** | - Histopathologically confirmed diagnosis of primary and brain metastases, - Provision of informed consent, - Willing to provide blood sample, | - Inability to meet all the inclusion criteria, - Diagnosed with or suspicion of synchronous or metachronous CNS malignancies, |

**Supplementary Table S11. Demographics of the Clinical Study-2 Cohort.**

This study determined the ability of the test to differentiate samples from GLI-M cases. NBT cases, cases of primary epithelial malignancy with brain metastases and healthy individuals (no diagnosis / suspicion of cancer).

|  | **Cohorts** | | | |
| --- | --- | --- | --- | --- |
|  | **GLI-M** | **NBT** | **EPI-M** | **Healthy** |
| **Total samples** | **40** | **22** | **24** | **500** |
| **Age**  Median  (Range) | 48  (2 – 73) | 47  (17 – 65) | 52  (25 87) | 43  (11 - 86) |
| **Gender**  Male  Female | 22  18 | 11  11 | 12  12 | 305  195 |
| **GLI-M**  *Astrocytoma, Gr II*  *Astrocytoma, Gr III*  *Ependymoma, Gr II*  *Ependymoma, Gr III*  *Glioblastoma, Gr IV*  *Glioma, NOS, Gr II*  *Glioma, NOS, Gr III*  *Oligodendroglioma, Gr II*  *Oligodendroglioma, Gr III* | 4  8  2  1  3  5  3  3  1 | - | - | - |
| **NBT**  *Arachnoid cyst*  *Benign Neuroparenchyma*  *Choroid Plexus Papilloma,*  *CP Angle Epidermoid Cyst*  *CP Angle Schwannoma*  *Meningioma*  *Pituitary Adenoma*  *Trigeminal Schwannoma*  *Ventricular Epidermoid Cyst* | - | 1  1  1  1  2  13  1  1  1 | - | - |
| **EPI-M**  *Breast*  *Colorectum*  *Esophagus*  *Esophagus+Breast*  *Stomach*  *Head and Neck*  *Kidney*  *Lung*  *Tested*  *Unknown Primary* | - | - | 7  2  2  1  1  2  1  6  1  1 | - |

**Supplementary Table S12. Clinical Study-2 Observations and Performance.** The table provides the summary of observations (status of CGCs and CTCs in each type of sample) as well as the summary of performance characteristics of the test in the Clinical Study-2.The Sensitivity, Specificity and Accuracy are based on the ability of the test to detect CGCs and differentiate GLI-M samples from all other types of samples.

|  | **Number** | **Post unblinding** | | | |
| --- | --- | --- | --- | --- | --- |
|  |  | **GLI-M** | **NBT** | **EPI-M** | **Healthy** |
| **Samples** | 586 | 40 | 22 | 24 | 500 |
| **CGC+, CTC+** | 0 | - | - | - | - |
| **CGC-, CTC-** | 522 | - | 22 | - | 500 |
| **CGC-, CTC+** | 24 | - | - | 24 | - |
| **CGC+, CTC-** | 40 | 40 | - | - | - |
| **Performance Characteristics** | | | | | |
| **Sensitivity** | - | 100%  (95%CI: 91.19% - 100%) | | | |
| **Specificity** | - | 100%  (95%CI: 99.33% - 100%) | | | |
| **Accuracy** | - | 100%  (95%CI: 99.37% - 100%) | | | |

**Supplementary Table S13. Inclusion-Exclusion Criteria for Clinical Study-3.**

| **Inclusion Criteria** | **Exclusion Criteria** |
| --- | --- |
| - Presenting with radiological ICSOL - No prior diagnosis of any cancers - Blood collection prior to tissue sampling - Availability of histopathological diagnosis - Provision of written informed consent - Willing to provide blood sample | - Inability to meet all the inclusion criteria, |

**Supplementary Table S14. Demographics of the Clinical Study-3 Cohort.** This prospective study determined the ability of the test to differentiate GLI-M and NBT in suspected cases with ICSOL.

|  | **GLI-M** | **NBT** | **Overall** |
| --- | --- | --- | --- |
| **Total samples** | **56** | **12** | **68** |
| **Age**  Median (Range) | 52 (6 – 77) | 58 (17 – 75) | 52 (6 – 77) |
| **Gender**  Male  Female | 36  20 | 6  6 | 42  26 |
| **GLI-M**  Astrocytoma, Gr II  Astrocytoma, Gr III  Glioblastoma, Gr IV  Glioma, NOS, Gr II  Glioma, NOS, Gr III  Oligodendroglioma, Gr II  Oligodendroglioma, Gr III | 6  9  30  1  1  1  8 | - | 6  9  30  1  1  1  8 |
| **NBT**  CP Angle Schwannoma  Craniopharyngioma  Meningiomas (Gr I)  Benign Neuroparenchyma  Benign Cystic lesion | - | 3  1  5  2  1 | 3  1  5  2  1 |

**Supplementary Table S15. Clinical Study-3 Observations.** The table provides the summary of observations (status of CGCs in each type of sample) in the Clinical Study-3.

| **Sample Type** | **Samples** | **Positive** | **Negative** |
| --- | --- | --- | --- |
| **GLI-M**  *Astrocytoma, Gr II*  *Astrocytoma, Gr III*  *Glioblastoma, Gr IV*  *Glioma NOS, Gr II*  *Glioma NOS, Gr III*  *Oligodendroglioma, Gr II*  *Oligodendroglioma, Gr III* | **56**  6  9  30  1  1  1  8 | **56 (100%)**  6  9  30  1  1  1  8 | **-**  -  -  -  -  -  -  - |
| **NBT**  *CP Angle Schwannoma*  *Craniopharyngioma*  *Cystic Lesion*  *Meningioma, Gr I*  *Benign Neuroparenchyma* | **12**  3  1  1  5  2 | **-**  -  -  -  -  - | **12 (100%)**  3  1  1  5  2 |

**Supplementary Table S16. Clinical Study-3 Performance.** The table summarizes the performance characteristics of the test in Clinical Study-3.

|  | **Number** | **Post unblinding** | |
| --- | --- | --- | --- |
|  |  | **NBT** | **GLI-M** |
| **Samples** | 68 | 12 | 56 |
| **Negative**  **(CGC-)** | 12 | 12 (100%) | - |
| **Positive**  **(CGC+)** | 56 | - | 56 (100%) |
| **Performance Characteristics** | | | |
| **Sensitivity** | - | 100%  (95%CI: 93.62% - 100% | |
| **Specificity** | - | 100%  (95%CI: 73.54% - 100%) | |
| **Accuracy** | - | 100%  (95%CI: 94.72% - 100%) | |

**Supplementary Table S17. Inclusion-Exclusion Criteria for Clinical Study-4.**

| **Inclusion Criteria** | **Exclusion Criteria** |
| --- | --- |
| - Presenting with radiological intra-axial SOL - No prior diagnosis of any cancers - Blood collection prior to tissue sampling - Availability of histopathological diagnosis - Provision of written informed consent - Willing to provide blood sample | - Inability to meet all the inclusion criteria, |

**Supplementary Table S18. Demographics of the Clinical Study-4 Cohort.** This prospective study determined the ability of the test to differentiate GLI-M, NBT and NGCM in suspected cases with intra-axial ICSOL.

|  | **GLI-M** | **NGCM** | **NBT** | **Overall** |
| --- | --- | --- | --- | --- |
| **Total samples** | **14** | **8** | **9** | **31** |
| **Age**  Median (Range) | 47 (14 - 72) | 28 (8 - 63) | 45 (19 - 65) | 45 (8 - 72) |
| **Gender**  Male  Female | 5  9 | 5  3 | 4  5 | 14  17 |
| **GLI-M**  *Astrocytoma, Gr III*  *Glioblastoma, Gr IV*  *Glioma, NOS, Gr III*  *Oligodendroglioma, Gr II*  *Oligodendroglioma, Gr III* | 2  8  1  1  2 | -  -  -  -  - | -  -  -  -  - | 2  8  1  1  2 |
| **NGCM**  *Medulloblastoma, Gr IV*  *CNS Lymphoma* | -  - | 5  3 | -  - | 5  3 |
| **NBT**  *Glial tumor, Low Grade*  *Pilocytic Astrocytoma, Gr 1*  *Hematoma*  *Ganglioglioma*  *Reactive Gliosis*  *Epidermoid cyst* | -  -  -  -  -  - | -  -  -  -  -  - | 3  1  2  1  1  1 | 3  1  2  1  1  1 |

**Supplementary Table S19 Clinical Study-4 Observations.** The table provides the summary of observations (status of CGCs in each type of sample) in the Clinical Study-4.

| **Sample Type** | **Samples** | **Positive** | **Negative** |
| --- | --- | --- | --- |
| **GLI-M**  *Astrocytoma, Gr III*  *Glioblastoma, Gr IV**  *Glioma NOS, Gr III*  *Oligodendroglioma, Gr II*  *Oligodendroglioma, Gr III* | **14**  2  8  1  1  2 | **13 (92.9%)**  2  7  1  1  2 | **1**  -  1  -  -  - |
| **NGCM**  *Medulloblastoma, Gr IV*  *CNS Lymphoma* | **8**  5  3 | **-**  -  - | **8 (100%)**  5  3 |
| **NBT**  *Glial tumor, Low Grade*  *Pilocytic Astrocytoma, Gr 1*  *Hematoma*  *Ganglioglioma*  *Reactive Gliosis*  *Epidermoid cyst* | **9**  3  1  2  1  1  1 | **-**  -  -  -  -  -  - | **9 (100%)**  3  1  2  1  1  1 |
| **includes a case of suspected Oligodendroglioma, Gr 3 / Glioblastoma, Gr 4.* | | | |

**Supplementary Table S20. Clinical Study-4 Performance.** The table summarizes the performance characteristics of the test in Clinical Study-4. The Sensitivity, Specificity and Accuracy are based on the ability of the test to detect CGCs and differentiate GLI-M samples from all other types of samples.

|  | **Number** | **Post unblinding** | | |
| --- | --- | --- | --- | --- |
|  |  | **NBT** | **NGCM** | **GLI-M** |
| **Samples** | 31 | 9 | 8 | 14 |
| **Negative**  **(CGC-)** | 18 | 9 (100%) | 8 (100%) | 1 (7.1%) |
| **Positive**  **(CGC+)** | 13 | - | - | 13 (92.9%) |
| **Performance Characteristics** | | | | |
| **Sensitivity** | - | 92.86%  (95%CI: 66.13% - 99.82%) | | |
| **Specificity** | - | 100%  (95%CI: 80.49% - 100%) | | |
| **Accuracy** | - | 96.77%  (95%CI: 83.30% - 99.92%) | | |
